# Study protocol for FAXAge: A randomized, controlled clinical trial of fasting and exercise to slow aging in humans

**DOI:** 10.64898/2026.02.28.26347327

**Authors:** Emma B. Fals, Emilie C. Springborg, Adam Bjørnholdt Berthelsen, Jonas Nyeman-Nielsen, Steen Larsen, Morten Scheibye-Knudsen

## Abstract

Biomarkers of aging, particularly DNA methylation-based clocks, have shown promise as tools to assess whether interventions may impact the rate of biological aging. Among possible interventions physical exercise has shown protective effects against many age-associated diseases, while time-restricted feeding (TRF), has shown metabolic benefits in preclinical models. The combined effect of exercise and TRF on aging biomarkers remains largely unexplored. In this 52-week four-armed, randomized, controlled trial (clinicaltrials.gov: NCT07207044) 240 healthy adults aged 65 and over will be allocated to four groups: combined cardio and strength training (EXE), TRF, combined EXE and TRF, or control. Participants will undergo assessments at baseline, 3, 6, and 12 months, with follow-ups at 2, 5, and 10 years. The primary outcome measure is DNA-methylation age with secondary measures including RNA-sequencing, metabolomics, inflammatory marker, microbiome analysis, cognitive and physical measures. By deeply phenotyping participants the Fasting And eXercise (FAXAge) study will provide novel insights into whether TRF, EXE, or a combination can slow or reverse biological aging in older adults.

## Background

Advances in medicine have among other factors led to an increase in global life expectancy across the 20^th^ and 21^st^ century^1^. In combination with declining birth rates it is projected that by the year 2050, one in every six people worldwide will be above the age of 65 years; in Europe, this number is already one in four people^2^. Although increasing life expectancy is positive, aging represents the largest risk factor for developing chronic diseases, and it has been suggested that aging *per se* should be regarded as a disease^3^. Indeed, aging interventions could impact not only lifespan, but also the multitude of diseases associated with old age e.g., T2D, cardiovascular disease and cancer^4^. An approach to impact all these chronic diseases could therefore be to treat aging itself thereby increasing both health- and lifespan.

Impacts of aging interventions in humans can be difficult to assess due to the long lifespan of humans. Efforts to find biomarkers of human aging that can be used to measure effects of human trials have therefore been explored for decades. About a decade ago, it was discovered that human aging tracks closely with changes in DNA methylation and subsequent work on these so-called aging clocks has shown that they are malleable for interventions ^5 6^. These biomarkers typically utilize machine learning methods to explore changes in multiple DNA methylation sites that appear to more strongly capture the complex changes in human aging compared to a single biomarker^7^. Importantly, this approach of using machine learning on more complex datasets from physiological to biochemical biomarkers has proven efficacious in capturing the often subtle changes that occur with age, for instance nuclear morphology can be used to predict cell senescence^8^, while biological age can be determined by DNA-methylation based aging clocks^7^ as well as portrait photos^8^. Still no consensus has been achieved on the composition of biomarkers for the best model for biological age and little is known about how the different biomarkers relate to each other and to physiological outcomes ^5^. Prospective intervention studies are therefore needed to establish and validate robust measures of biological age to build on the recent advances in the field^9^.

Being physically active is associated with increased life expectancy, potentially by attenuating age-related physiological alterations^10^. These include the preservation of a healthier body composition with reduced total and abdominal fat, greater skeletal muscle mass, and higher bone mineral density, as well as improved cardiovascular function, enhanced insulin sensitivity, and lower levels of systemic inflammatory biomarkers^10^. Collectively these adaptations contribute to the protective effects of exercise against most chronic age-related diseases^11^.

Conversely, intermittent fasting and caloric restriction have been shown to increase lifespan in organisms from mice to rhesus monkeys^12–14^ and improve health parameters in humans^14^. TRF is a type of intermittent fasting in which no calories are consumed for a period, typically 12-18 hours per day. It is currently unknown whether this type of sustainable fasting improves markers of aging in older adults, although data suggests that periods of fasting can be healthy^14^. For instance, in rodents, TRF has shown protective effects against metabolic dysfunctions induced by the Western diet such as obesity, cardiovascular disease, hypertension, diabetes^14^. These benefits include reduction in body weight, increase in energy expenditure, improved glycemic control, and lower insulin levels, as well as decreases in hepatic fat and hyperglycemia^14^. The metabolic protection after TRF has been proposed to be due to metabolic adaptations, characterized by increased amounts of circulating ketones, while fatty acids, glucose and insulin remain at low levels^14^. Importantly, TRF has been shown to confer the major lifespan benefit of a caloric restricted diet in mice^15^At the cellular level, it is believed that some of the effects of fasting are mediated through the activation of AMP-activated protein kinase (AMPK), recycling of cellular components via autophagy, reduced mTOR activation and increased mitochondrial biogenesis ^14^. However, loss of muscle and bone-mass may limit the use of this intervention in older adults where this could be a serious side effect^16^. Fasting combined with exercise could be a mean to mitigate the possible catabolic effect of intermittent fasting^17^.

To address these issues, this case-control clinical trial will investigate biological age and physiological and molecular outcomes in an intervention setting with long-term follow up. This may allow us to answer the following key questions which are critically needed for the field: 1) Which markers of biological age are clinically relevant? 2) Is biological age amenable for interventions? 3) Which biological age measurement is most reproducible? 4) Is biological age predictive of health and future outcomes? 5) Are the non-invasive biomarkers as good as the more invasive biomarkers? 6) How do different biomarkers associate with each other and other health parameters? To answer these questions, a 52 weeks four-armed randomized, controlled intervention trial will be conducted with subjects randomized to either a combined cardio and strength exercise (EXE), fasting via time-restricted feeding (TRF), combined TRF and EXE (FAX), or control group (Figure 1). This trial will provide new insight into whether TRF and exercise may reduce or potentially reverse the rate of human aging.

**Figure 1.**
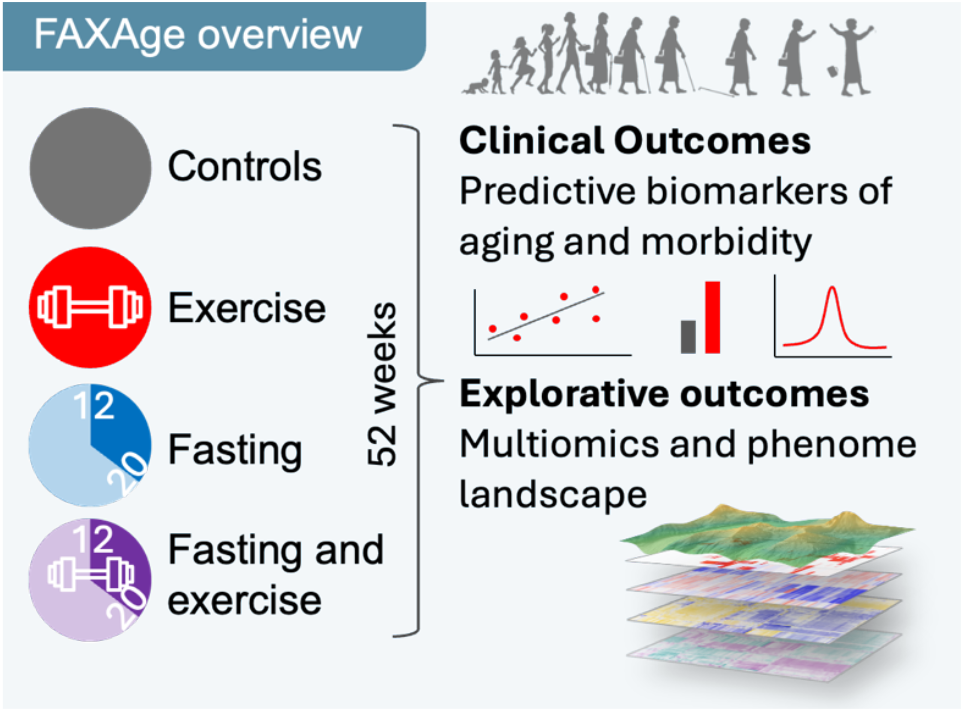
Overview of the study.

We hypothesize that exercise, intermittent fasting and a combination of the two will provide similar, superior benefits to DNA methylation epigenetic age when compared to controls after 52 weeks. It is furthermore hypothesized that the interventions will provide similar, superior benefits to CRP, TNF-α, IL-6, IL-8, NAD+, hematologic age, cell senescence in blood smear, cognitive function, transcriptional age (RNA-sequencing), metabolomic changes, microbiome changes, functional age (handgrip strength, gait speed, VO_2_max), body composition, vocal age, and photo age when compared to controls after 52 weeks.

## Methods

### Study design

The study visits will be conducted at the Department of Biomedical Sciences, University of Copenhagen. 240 healthy elderly aged 65 and older will be recruited and block randomized to either of the interventions for 52 weeks. Assessments are carried out at baseline, 3 months, 6 months, and 12 months and followed up after 2 years, 5 years and 10 years (Figure 2).

**Figure 2.**
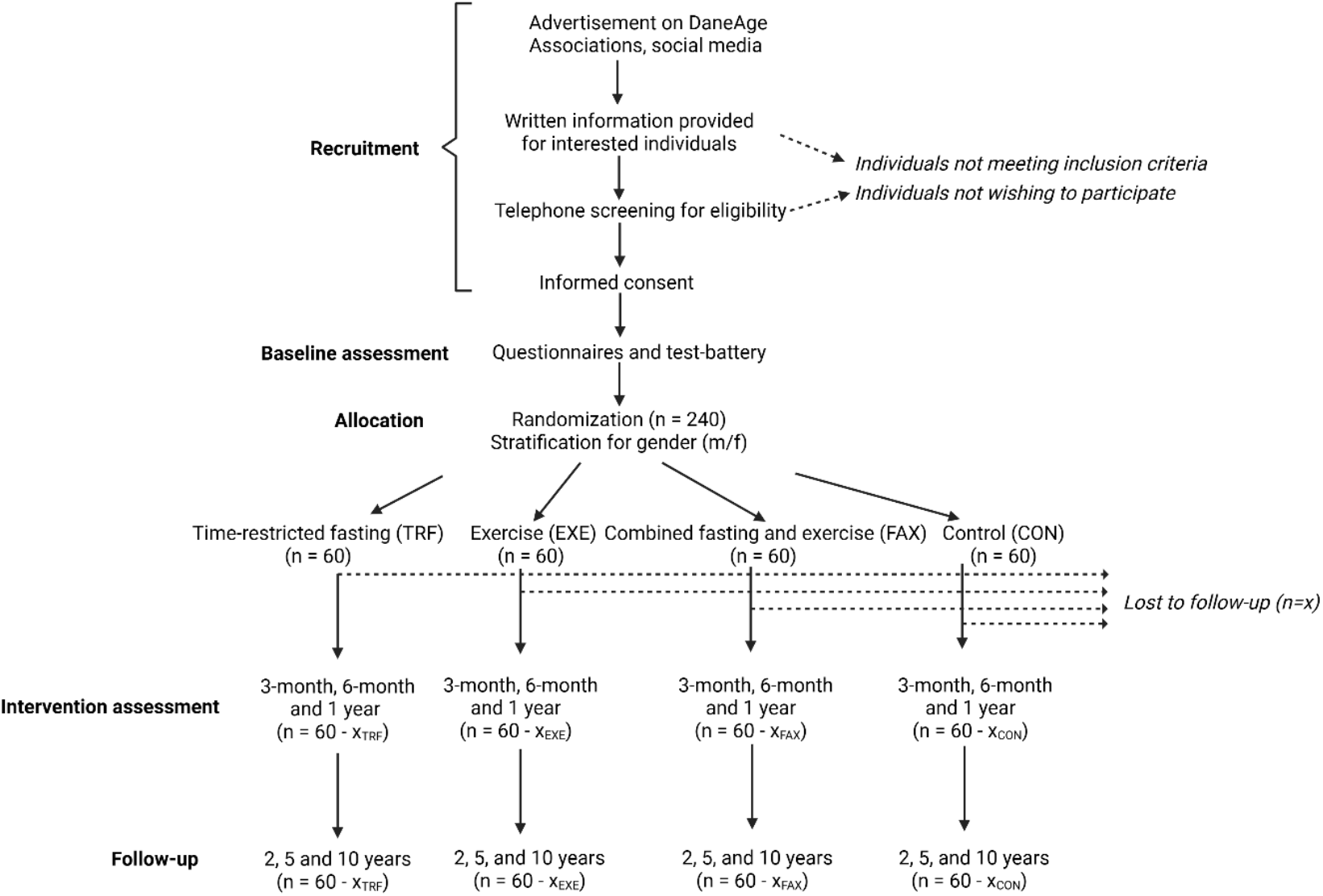
Flow chart of the FAXAge experimental set-up.

### Participants, Recruitment Strategy, and Randomization

Participants are excluded if they have received another investigational drug or intervention within 1 year, as prior treatments could influence the responses observed in this study, making it difficult to isolate effects to the current intervention. Individuals engaging in more than one hour of systematic strenuous exercise or strength training a week are excluded to maintain similar baseline activity levels across participants and reduce variability in fitness, making the effects of the intervention clearer. Individuals already practicing time-restricted eating are excluded, as they will not show clear pre- and post-intervention responses. We wish to include a representative part of the healthy elderly population, however, we expect significant bias in our data if we only include individuals without chronic diseases. For this reason individuals with severe or dysregulated diseases are excluded, as the disease state may independently impact the intervention outcomes, while allowing one well-managed chronic disease helps limit heterogeneity and risks for the participants. Smoking is an exclusion criterion due to its effects on cardiometabolic health, inflammation, and exercise capacity, all of which could confound study results. Finally, individuals using systemic glucocorticoids, androgens, or antiandrogens are excluded because these medications have strong effects on metabolism, muscle adaptation, appetite, and glucose tolerance, potentially masking or exaggerating the effects of time-restricted feeding and exercise interventions.

Participants are recruited via online advertising, on social media, and in the DaneAge Association membership magazine.

Participants will be randomized according to a permuted block randomization scheme with a block size of 4, stratified according to biological sex. The randomization list will be generated prior to commencing the study.

### Interventions

Participants randomized to the EXE and FAX group will perform supervised resistance training two times per week as well as cardio training at home two times per week monitored by fitness watches to track heart rate. This combined approach is chosen because both high muscle strength and cardiorespiratory fitness are independently associated with lower all-cause mortality, whereas low levels correlate with increased risk^18^. Accordingly, this intervention is designed to enhance both muscle strength and cardiorespiratory fitness, enabling the investigation of their association with changes in markers of biological age.

The strength training intensity will be individualized and increased over time according to changes in the participant’s training status, aiming to maintain a similar relative load across participants by adjusting loads to a target range of repetitions in reserve. The training program will consist of the same exercises over the 52 weeks and is designed in a progressive manner as this approach has been shown to enable long-term adaptations in muscular strength through progressive increases in mechanical loading and total training stimulus, thereby limiting training plateaus over time^19^. It will consist of approximately 1 hour of strength training per session with at least 48 hours between each session. Participants will perform a standardized, machine-based (Technogym S.p.A, Cesena, Italy) strength training program involving exercises for the lower and upper body. The program includes both multi-joint (leg press, chest press, lateral pulldown and low row) and single-joint (leg extension and lying leg curl) exercises, with an emphasis on multi-joint movements, which have been shown to elicit greater improvements in muscle strength and contribute to improvements in cardiorespiratory fitness compared with single-joints exercises when total training load is matched^20^This effect has been attributed to a higher oxygen demand associated with the involvement of a larger active muscle mass ^20^.

The training program is initiated with a 4-week familiarization phase characterized by high-repetition, low-load training, to ensure proper technique, allow musculoskeletal tissue adaptation and to reduce the risk of injury^21^. The training period consists of 4 blocks of 13 weeks. Each block begins with three sets of 12 repetitions per exercise, with the number of repetitions decreasing in a stepwise manner every three weeks, ultimately concluding with four sets of six repetitions in the last three weeks. Within each block, training will be organized into consecutive 3-week phases during which repetition range, target repetitions in reserve, and number of sets were maintained constant, allowing sufficient time for adaptation before progressing to the next phase. After completion of each block, the same progression pattern is repeated in the subsequent block with adjusted absolute loads. Every 13th week a deload phase is implemented to facilitate recovery and mitigate fatigue development^22^. During this week, training volume is reduced by ∼30-50% by reducing the number of sets for each exercise and increasing repetitions in reservewhile maintaining the target repetition range. Training sessions will be supervised and will take place at the Department of Biomedical Sciences, University of Copenhagen.

Cardio training will be of self-chosen modality and at moderate to vigorous intensity (>70% HRmax) since vigorous-intensity exercise may result in greater increases in aerobic capacity than moderate-intensity^23^ and that participation in vigorous activities is associated with lower mortality rates^24^. The duration will increase from 30 minutes per session in the first month, 45 minutes per session in the second month to 60 minutes per session in the following 3-12 months. Adherence, workout duration, and average and maximal heart rate will be monitored by fitness watches provided to each participant.

Participants randomized to the TRF group will be instructed to abstain from any caloric intake during the targeted fasting window of 16 continuous hours and consume ad libitum during the 8-hour eating window. Participants can choose their preferred eating window and are encouraged to drink plenty of water during fasting. Participants will receive an adherence diary including an eating time log for noting the time of first and last calorie consumption each day.

Participants randomized to the FAX group will be instructed to both exercise and fast during the 52-week intervention.

### Measurements

Participants go through a battery of tests before the intervention (Figure 3), 3 months and 6 months into the intervention and after the intervention. Additionally, follow-up testing is performed at 2, 5 and 10 years after the intervention. Follow-up testing is deemed critical because it may allow us to discover long term beneficial or detrimental effects of these interventions. In the section below we will go through the different data modalities that will be gathered as part of the intervention and the rational for why these measures were chosen. Tests are performed according to defined standard operating procedures (SOPs) in the same order for all participants at all visits, and to ensure a minimum of bias, test leaders are instructed to not look at the participants’ previous test results before a new test.

**Figure 3.**
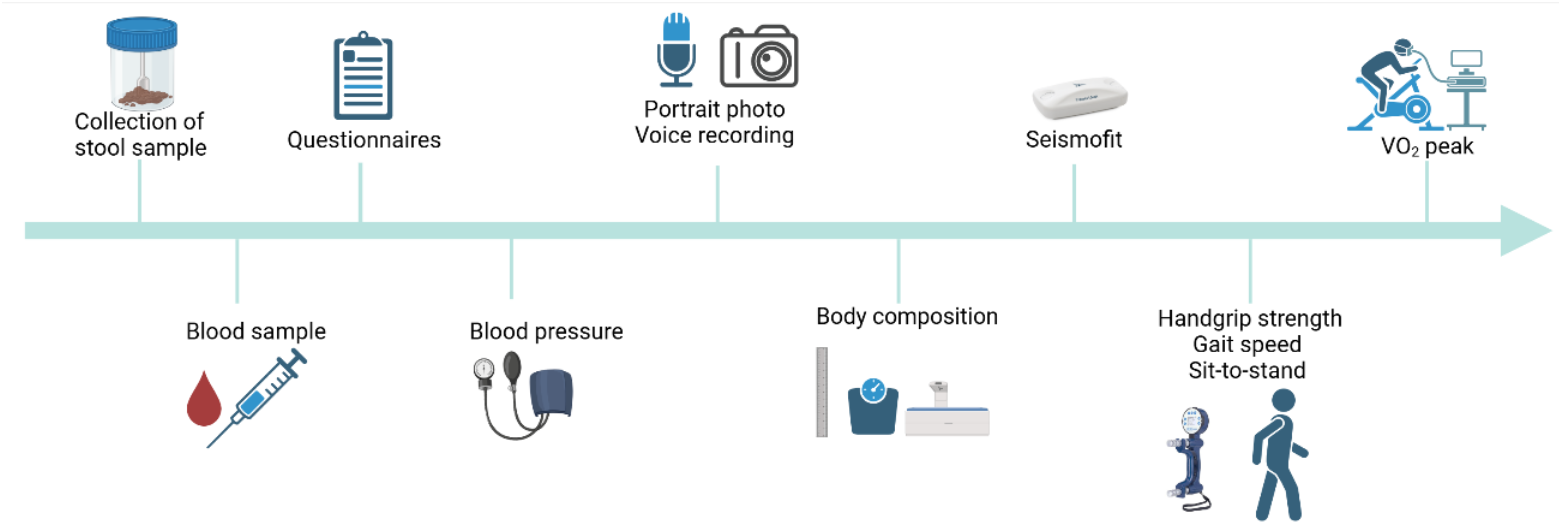
Visualization of the test-battery. Participants will be tested at baseline, after 3 months, 6 months and 1 year and follow-up will be performed 2, 5 and 10 years after baseline. A stool sample is collected for microbiome analysis, then a blood sample is taken to assess multiple blood markers, participants fill out questionnaires, have their blood pressure taken and digital xx are taken. Then body composition is measured with a DXA-scan and VO_2_peak is estimated with a Seismofit before physical tests are performed. Participants perform handgrip strength, gait speed and sit-to-stand tests as well as a graded cardiopulmonary VO_2_peak test.

#### Stool sample

Changes in the gut microbiome has been seen with age and may predict health outcomes in the elderly^25–27^. Notably, gut microbiome is strongly affected by feeding habits and we expect TRF will have a considerable effect on this^28^. For that reason, all participants will be given a feces kit to collect a stool sample prior to the first visit. The stool sample will be collected on the test day and stored at -80 degrees Celsius until analysis.

#### Blood work

Venous blood is collected following a 12-hour overnight fast at the start of the test day and used for analysis of multiple parameters (Table 1). Measurement of inflammatory markers is important, as chronic, low-grade inflammation is a hallmark of aging and has been linked to an increased risk of frailty and mortality^29–31^. Glycemic control markers are measured because impaired glucose regulation is common among older adults^32^ and is associated with elevated risk of most chronic age-associated diseases ^33^. Complete blood cell count parameters are assessed since aging influences hematopoiesis, and white blood cell counts serve as predictors of all-cause mortality^34^.

**Table 1.** Measured blood markers.

| Category | Blood marker |
| --- | --- |
| Inflammatory markers | TNF-α, IL-6, IL-8, CRP |
| Glycemic control | HbA1C, Glucose |
| Hematology | Hb, Erythrocytes, Hct, MCH, MCHC, MCV, Platelets, RBC, Leukocytes, Basophils, Eosinophils, Monocytes, Lymphocytes, Neutrophils |
| Molecular tests | DNA methylation, RNA-sequencing, Metabolomics |
| Biochemical | ALAT, ASAT, Alkaline phosphatase, Albumin, Urea, Total protein, Iron, Bilirubin, Lactate acid dehydrogenase, Calcium, Creatinine, Total cholesterol, HDL, LDL, Triglycerides, Selenium, Q10, NAD<sup>+</sup>, Sodium, Potassium, Chloride |

**Table 2.** List of Outcomes.

| Procedure | Baseline | 3 months | 6 months | 1 year | Follow-up<br>(2, 5, 10 years) |
| --- | --- | --- | --- | --- | --- |
| DNA methylation (primary) | X | X | X | X | X |
| Questionnaire | X | X | X | X | X |
| MoCA | X | X | X | X | X |
| Height | X | X | X | X | X |
| Weight | X | X | X | X | X |
| Body composition | X | X | X | X | X |
| Blood pressure | X | X | X | X | X |
| Blood samples | X | X | X | X | X |
| Microbiome sequencing | X | X | X | X | X |
| Handgrip strength | X | X | X | X | X |
| Gait speed | X | X | X | X | X |
| Seisomfit | X | X | X | X | X |
| VO<sub>2</sub>peak | X | X | X | X | X |
| Voice recording | X | X | X | X | X |
| Portrait photo | X | X | X | X | X |

DNA methylation analysis is performed to evaluate epigenetic changes and biological age, while RNA-sequencing provides insights into transcriptional, translational, and signaling processes at the cellular level. Metabolomic profiling will be performed on plasma to assess the changes, exercise and fasting induce at the molecular level and what adaptations and cellular pathways are involved. Lastly, biochemical and metabolic markers are measured to detect organ system decline and age-related metabolic disturbances. Combined with physiological measures this will provide a comprehensive understanding of how fasting and exercise influence aging.

To explore classic aging clocks, DNA-methylation analyses will be performed on peripheral blood mononucleated cells. We choose to isolate these cells instead of performing investigations on whole blood to limit the potential effect of blood composition changes that may be a side effect of interventions. A finger prick is also performed on the tip of the middle finger for thin blood smear senescence prediction^35^.

#### Questionnaires

To gain understanding of overall health, sleep quality, and cognitive decline participants answer the following questionnaires:

##### Health Examination

Participants will answer a modified version of the Danish Health Examination Survey questionnaire, including questions about chronic diseases, dietary habits, alcohol, stress, physical activity and for female participants their menstrual cycle (see appendix).

##### Sleep Quality

Pittsburgh Sleep Quality Index (PSQI) questionnaire will be used to assess subjective sleep quality and disturbances over the past month. Questions include assessment of duration, latency, efficiency, disturbances, and day-time dysfunction. Sleep will be further evaluated through wearables.

##### Montreal Cognitive Assessment (MoCA)

The MoCA Full test is performed to assess a wide range of cognitive domains, including memory, attention, executive function, language, visuospatial abilities and orientation, to possibly identify mild cognitive impairment (MCI). This allows for identification of subtle cognitive deficits and is better suited in healthy elderly than the mini-mental state examination^36^.

#### Vitals

Participants’ resting pulse and blood pressure is measured three times on the left arm using an electric blood pressure monitor in a seated position with both feet flat on the floor and the arm resting on the table. Blood pressure is measured after filling out questionnaires to ensure that the participant is relaxed. Three consecutive readings are obtained automatically, and the mean of the values is used. If the variability between the first three consecutive readings exceeds threshold, the blood pressure measurement is repeated and the outlier is discarded^37^.

#### Portrait photo and voice recording

A portrait photo as well as a voice recording, answering a standardized question, will be obtained of each participant. Audio clips are expected to be 1 minute. Both will be used for determination of the participants’ biological age by using facial image-based and voice-based age prediction algorithms using deep neural networks trained to predict age ^8,38^

#### Body composition

Body composition and bone mineral density are evaluated with dual energy X-ray absorptiometry (DEXA)-scan (Lunar iDEXA, Madison, Wisconsin, USA) using enCORE software, V.18^39^. A whole-body scan is performed to determine fat-percentage, lean body mass and bone mineral density.

#### Handgrip strength

Grip strength is measured to assess the maximal isometric handgrip strength in kilograms with a Jamar Smart Hand Dynamometer (Jamar, Nottinghamshire, UK). Participants are seated in a chair with a straight back with the elbow bent in a 90-degree angle. The dynamometer is pointing vertically upwards. The measurement is a 5 second maximum-effort measure and is repeated three times on each hand with one minute rest in between trials on the same hand. The participants are verbally encouraged during each maximum-effort measure. The highest value obtained for each arm is reported and used for data analysis.

#### Gait speed

Participants’ gait speed and function is assessed by a standardized 4-meter test, where participants are instructed to walk two times – once at their normal pace and once as fast as they can. Participants are also video recorded while walking and gait speed, acceleration, balance, cadence, step length, posture and joint angles will be estimated using Tracked Biotechnologies’ TrackedGait system (Tracked Biotechnologies, Virginia, USA).

#### 30 second sit-to-stand

A 30 second sit-to-stand test is performed to assess strength in the lower extremities. The test is performed using a chair (45 cm) without armrest. Participants are shown how to perform the test and instructed to place their feet flat on the ground, cross the arms and hold them against the chest. Participants must stand fully and contact the seat fully. The score is the number of full stands completed within 30 seconds.

#### VO_2_peak

##### Estimated VO_2_peak

VO_2_peak will be estimated at rest using seismocardiographic measurements (Ventriject, Copenhagen, Denmark)^40^. This will be done immediately after the DEXA scan where participants are lying down and are relaxed.

##### Direct measure

A cardiopulmonary exercise test (CPET) is performed on a Monark LC7TT bike (Monark, Varberg, Sweeden) using the Quark CPET metabolic cart (Cosmed, Rome, Italy) to determine the peak oxygen consumption rate using indirect calorimetry. Participants perform a graded exercise test with warm-up and increment intensity adjusted to their Seismofit-estimated VO_2_. Estimated peak power output is calculated from the estimated VO_2_peak, a sex-specific coefficient, body weight and a regression-based constant interpreted^41^. From estimated peak power output, warm-up intensity and increment size is calculated to reach a total test duration of 10-12 minutes including a 5-minute warm up^42^.

#### Activity monitors

Activity monitors (Huawei Band 10, Huawei, Shenzhen, China) will be provided to all participants to monitor daily activity. The activity monitors will be used by EXE and FAX participants to monitor home workouts of self-chosen modality and is used to ensure that average heart rate is >70% HRmax. For CON and TRF the activity monitors are used to assess baseline activity level. Furthermore, the monitors are used to assess sleep quality, which will be compared to the PSQI scores.

## Retention

### Biweekly phone calls

To increase participant retention and compliance with the randomly prescribed interventions, personalized attention will be provided through biweekly phone calls throughout the intervention for all four groups. This will give the opportunity to discuss individual challenges and tailor solutions to help participants adhere to the various interventions. However, there will not be applied pressure on individuals to continue their enrollment in the trial if they wish to be removed from the trial.

### Strategy for follow-up assessment

Participants will be reminded by email ∼2 months prior to follow-up assessments. The email will be followed up by a phone call shortly after to schedule a date for the follow-up test day. An email with confirmation of the appointment will be sent right after the phone call. A reminder email confirming the agreed time and date will be sent out ∼ 2 weeks prior to the follow-up as well as a reminder email with practical information the day before the test day.

## Statistical considerations

Sample size determination for the interventions trial is conducted using the primary outcome DNA methylation measured at 52 weeks for the pooled three intervention groups (EXE, TRF, FAX) versus the control group. Biological age prediction using epigenetics yields a standard deviation of ∼10% ^43^. Assuming an expected pooled effect size for the intervention groups versus the control group is considerable (Cohen’s d = 1.0). With a probability of a Type I and Type II error set at α = 0.05 and β = 0.20, respectively, the estimated sample size was 40 participants allocated in a 1:3 ratio (controls:intervention groups). Our 3-month pilot study (Slowage, clinical trials id: NCT05593939) as well as a similar previous 1-year training intervention study with older individuals showed low dropout rates around 5-10% ^44^. To accommodate the possibility of a higher dropout rate, a total of 240 participants will be recruited whereby 60 participants will be included in each of the four groups (EXE, TRF, FAX, CON), allowing for a 33% dropout rate. Data from dropouts will not be included in the primary outcomes but may be used in exploratory outcomes.

## Personal information

The identities of all participants will be known and stored at the laboratory site at the University of Copenhagen (UCPH). This information will not include the participants’ contact details and identification details. Instead, each participant and their associated research data will be identified by a unique study identification number. Data will be stored and managed in full compliance with regulatory bodies and according to GDPR. Study data will be managed on the secure, web-based software platform REDCap where it will be secured and password-protected and only accessible for research staff working on the project. At the conclusion of the study, all study databases will be de-identified and archived at UCPH.

## Discussion

The FAXAge study investigates the combined and individual effects of physical activity and TRF in a year-long randomized intervention trial. To our knowledge, no previous study has combined these two interventions in older adults, potentially providing novel insights into how dietary manipulation and structured exercise interact to influence the rate of aging. While the role of physical activity in promoting healthy aging is well supported by research and clinical evidence^45–47^, little is known regarding the effects of exercise on rates of aging. Further, while work in preclinical models indicate that TRF is among the strongest influencers of aging and lifespan in animals it is unknown if sustained intermittent fasting interventions can influence health and rates of aging in humans.

This trial will provide important evidence on the effects of intermittent fasting and exercise, both independently and in combination, on molecular biomarkers of biological aging in older adults. The use of multiple measures of biological aging, including molecular, physiological, and digital biomarkers, will enable a multidimensional assessment of aging processes. This integration of lifestyle interventions and biomarkers will address a knowledge gap regarding whether the rate of aging can be reduced or potentially slowed through lifestyle interventions, an important finding that could inform health policies for societies across the globe.

Should TRF, exercise, or the combination prove effective, these strategies could represent affordable, scalable, and safe strategies to promote healthy aging. This intervention study could potentially shape governmental lifestyle recommendations for healthy aging, as such interventions may reduce the prevalence of age-related diseases like cardiovascular disease, diabetes, and cognitive decline, thereby promoting independence among older adults. Enhanced physical function and metabolic health also reduce the frailty and fall risk, leading to fewer hospitalizations and lower care needs^48,49^.

Beyond health benefits, these interventions have economic implications. Given they lower disease burden and subsequently the healthcare utilization, they reduce the financial burden on health systems and extend the productive lifespan of older adults. Research suggests that an increase in life expectancy by 1 year is worth 38 trillion US dollars, while an increase in life expectancy by 10 years is worth 367 trillion US dollars^50^. Promoting fasting and exercise in elderly populations, therefore, holds potential to generate significant economic returns.

Additionally, improved physical and cognitive health supports greater social participation and community engagement among older adults, which in turn enhances quality of life^51^. Indeed, societal participation may not only increase perceived health but also reduce mortality, cognitive disability, frailty, and prevalence of depression^51^. Thus, giving older adults the guidelines for healthy aging, this intervention could empower them to contribute to society, benefiting both their quality of life, society and the economy. Furthermore, healthy aging may encourage older adults to reenter or remain in the workforce, potentially increasing annual GDP, thus benefiting the economy^52^.

Follow-up assessments are performed after 2, 5 and 10 years which will also include questionnaires regarding behavior such as food and exercise. This will allow assessment of whether the potential beneficial effects of the interventions are sustained over time after the intervention ends. Furthermore, it will identify potential delayed or long-term benefits or adverse effects that might not be evident immediately after the intervention. Follow-ups will provide data on how fasting and exercise influence outcomes like disease risk and mortality in the elderly.

## Ethics approval

The FAXAge study is registered at Clinicaltrials.gov (NCT07207044) and has been approved by the regional ethical committee (Capital Region, Copenhagen, No. H-25020642) and complies with the declaration of Helsinki. Oral and written information is provided, and oral and written consent will be obtained before onset of interventions. Participation is voluntary and participants can terminate their study participation at any time. Changes or additions to the protocol are not implemented before approval by the ethics committee.

## Author contributions

EBF and ECS wrote and edited the manuscript

ABB and JNN wrote the section on the exercise intervention

SL and MSK supervised and edited the manuscript

## Data Availability Statement

Not applicable

